# Limited durability of protection conferred by XBB.1.5 vaccines against Omicron-associated severe outcomes among community-dwelling adults aged ≥50 years, Ontario, Canada, September 2023 to June 2024

**DOI:** 10.1101/2024.12.14.24318807

**Authors:** Nelson Lee, Lena Nguyen, Sharifa Nasreen, Peter C. Austin, Kevin A. Brown, Sarah A Buchan, Ramandip Grewal, Kevin L Schwartz, Mina Tadrous, Kumanan Wilson, Sarah E Wilson, Jeffrey C Kwong, the Canadian Immunization Research Network (CIRN) investigators

## Abstract

We estimated XBB.1.5 vaccine effectiveness against hospitalization/death among adults aged ≥50 years. Compared with non-XBB.1.5 vaccinees, the initial protection of 64% (95%CI, 57%−69%) was reduced when JN/KP-sublineages became predominant, and quickly declined. No significant protection was observed >6 months post-vaccination. Short durability of protection poses unique challenges for COVID-19 vaccination.

## BACKGROUND

Omicron became the globally predominant circulating SARS-CoV-2 variant in early 2022, and has since evolved into successive, immune-evasive sublineages. We previously reported declining effectiveness of booster doses of original monovalent vaccines against severe Omicron-associated outcomes during periods of BA.1/BA.2, BA.4/BA.5, and BQ/XBB predominance [1], which was only partially boosted by Omicron-containing bivalent vaccines [2]. The XBB.1.5 monovalent vaccine became available in Ontario, Canada in fall 2023 to individuals aged ≥6 months to provide protection against the then-predominant sublineage; higher-risk persons (i.e., those aged ≥65 years, moderately/severely immunocompromised individuals, and long-term care residents) were subsequently recommended to receive an additional dose in spring 2024 [3,4]. In this study, we aimed to estimate the relative effectiveness and durability of protection conferred by mRNA XBB.1.5 vaccines compared with prior vaccines over the period from September 2023 (launch of the monovalent XBB.1.5 vaccination program in Ontario) to June 2024, during which the XBB sublineage was replaced by JN/KP sublineages.

## METHODS

We conducted a population-based, test-negative design study using linked databases in Ontario to estimate effectiveness of mRNA (Pfizer or Moderna) XBB.1.5 vaccines against Omicron-associated severe outcomes (hospitalization or death) over time. The study population included community-dwelling adults aged ≥50 years who underwent ≥1 polymerase chain reaction (PCR) test for SARS-CoV-2 from September 24, 2023 to June 1, 2024. We excluded individuals who received an incomplete series (i.e., only 1 dose), or non-Health Canada-authorized or non-mRNA vaccines for their primary series (n=536); were immunocompromised (n=12,332); or tested positive within the past 60 days (n=99).

As described in our previous studies [1,2], we used province-wide data from ICES, an independent, non-profit research institute whose legal status under Ontario’s health information privacy law allows it to collect and analyze health care and demographic data for health system evaluation and improvement. SARS-CoV-2 laboratory testing, COVID-19 cases surveillance, COVID-19 vaccination, and health administrative datasets were linked using unique encoded identifiers and analyzed at ICES. Data on rapid antigen tests were unavailable.

We defined cases as Omicron-associated hospitalization or death attributable to SARS-CoV-2 infection confirmed by PCR. We defined controls as individuals who were symptomatic and tested negative for SARS-CoV-2 by PCR, but were not necessarily hospitalized. A sublineage-predominant period was defined as ≥50% of sequenced samples being confirmed with a specific Omicron sublineage. In Ontario, the predominant sublineages were identified as XBB (24 September 2023−23 December 2023) and JN/KP (24 December 2023−1 June 2024) (**Supplementary Figure S1**). Data on individual-level whole genome sequencing were unavailable.

In the primary analysis, we used multivariable logistic regression to estimate the relative change in the odds of severe outcomes (defined as the proportionate reduction in the odds of hospitalization/death) associated with XBB.1.5 vaccination, compared to subjects who had received ≥2 doses of non-XBB.1.5 vaccines (i.e., original monovalent or Omicron-containing bivalent vaccine), with the last dose ≥6 months prior to the date of SARS-CoV-2 testing. Relative vaccine effectiveness (rVE) was calculated using the formula (1−adjusted odds ratio)*100%, and was reported separately for XBB-predominant and JN/KP-predominant periods, and at 0−<3 months, 3−<6 months, and 6−<9 months since XBB.1.5 vaccination. The models were adjusted for age, sex, public health unit region, neighborhood-level socio-demographic variables, influenza vaccination status (proxy for health behaviors), comorbidities, receipt of home care services, and week of test (modeled using restricted cubic splines) [1,2].

In secondary analyses, we estimated rVE of XBB.1.5 vaccine stratified by: (1) age group (≥65 years *vs*. 50−64 years); and (2) the type of non-XBB.1.5 vaccine received most recently (original monovalent *vs*. Omicron-containing bivalent vaccine). We conducted a sensitivity analysis to examine the initial (0−<3 months) benefit of XBB.1.5 vaccine compared with variable lengths of time since the last non-XBB.1.5 vaccine (≥12, 6−11, or <6 months prior). We also estimated absolute VE by comparing to subjects who remained unvaccinated (n=2,060, <10% of the entire cohort).

We used SAS version 9.1 (SAS Institute Inc., Cary, NC) for all analyses. All tests were 2-sided, with a p-value <0.05 indicating statistical significance.

## RESULTS

We included 4,895 cases with Omicron-associated severe outcomes and 23,223 test-negative controls (among a total of 24,498 unique individuals), with 11,287 and 16,831 subjects tested during the XBB- and JN/KP-predominant periods, respectively [**Supplementary Table S1**]. For both periods, cases were older than controls (e.g., median age 81 [IQR 73−88] years *vs*. 73 [IQR 62−83] years; and 81 [IQR 73−88] years *vs*. 74 [IQR 63−84] years, during the respective periods), and higher proportions were male or had comorbid conditions, while fewer had prior documented PCR-confirmed SARS-CoV-2 infections (12.4% *vs*. 26.3% and 13.7% *vs*. 21.6%, respectively). Notably, we found no substantial differences between cases and controls in their prior vaccination history (unvaccinated *vs*. receipt of original monovalent or Omicron-containing bivalent vaccines [i.e., non-XBB.1.5 vaccines]).

During the XBB-predominant period, 1,686 subjects had received XBB.1.5 vaccines, at a median interval of 27 (IQR 13−42) days before the date of testing; whereas 8,814 had only received non-XBB.1.5 vaccines, at a median interval of 511 (IQR 348−684) days prior. During the JN/KP-predominant period, 6,070 had received XBB.1.5 vaccines and 9,488 had received only non-XBB.1.5 vaccines, at median intervals of 110 (IQR 73−154) days and 743 (IQR 532−841) days prior, respectively.

During the XBB-predominant period, compared to those who only received non-XBB.1.5 vaccines with the most recent dose ≥6 months ago, rVE of XBB.1.5 vaccination was 64% (95%CI, 57%−69%) after 0−<3 months (**Figure 1**). During the JN/KP-predominant period, rVE was 57% (95%CI, 48%−64%) after 0−<3 months, and declined over time to 44% (95%CI, 32%−54%) after 3−<6 months and 21% (95%CI, −15% to 46%) after 6−<9 months.

**Figure 1.**
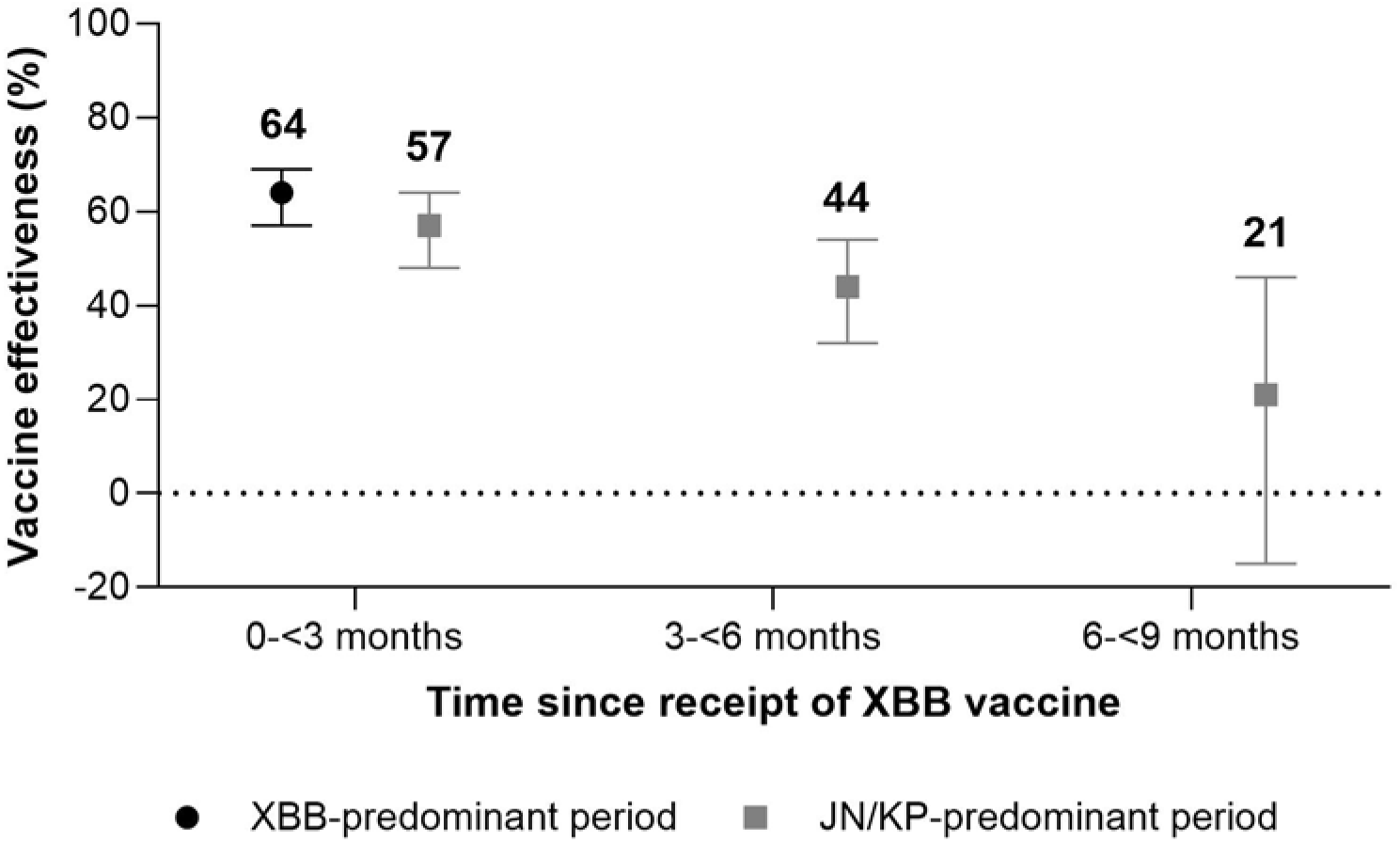
Relative vaccine effectiveness of mRNA XBB.1.5 vaccination over time, compared to subjects who received only non-XBB.1.5 vaccines at least 6 months prior, during periods of XBB and JN/KP sublineage predominance.

Stratified by age group, rVE after 0−<3 months was 68% (95%CI, 62%−73%) and 56% (95%CI, 25%−74%) for subjects aged ≥65 and 50−64 years, respectively, during the XBB-predominant period (**Supplementary Figure S2**); rVE after 0−<3 months was 60% (95%CI, 52%−67%) and 58% (95%CI, 14%−80%), respectively, during the JN/KP-predominant period. Stratified by the type of non-XBB.1.5 vaccine last received by the reference group, rVE of XBB.1.5 vaccination after 0−<3 months was 71% (95%CI, 66%−76%) and 60% (95%CI, 53%−67%) relative to recipients of the original monovalent and bivalent vaccines, respectively, during the XBB-predominant period (**Supplementary Figure S3**), whereas rVE was 62% (95%CI, 54%−68%) and 47% (95%CI, 35%−57%) after 0−<3 months, and 53% (95%CI, 42%−62%) and 43% (95%CI, 28%−55%) after 3−<6 months, respectively, during the JN/KP-predominant period. rVE estimates were not significant beyond 6 months, irrespective of the type of non-XBB.1.5 vaccine last received.

In the sensitivity analysis, rVE of XBB.1.5 vaccination was higher when the reference group received a non-XBB.1.5 vaccine more remotely compared to more recently (e.g., during JN/KP predominance, rVE was 59% [95%CI, 51%−66%] compared to subjects last vaccinated ≥12 months prior, versus 26% [95%CI, −7% to 49%] compared to subjects last vaccinated 6−11 months prior) (**Supplementary Figure S4**). Absolute VE at 0−<3 months after XBB.1.5 vaccination was 73% (95%CI, 64%−80%) during the XBB-predominant period and 67% (95%CI, 53%−76%) during the JN/KP-predominant period.

## DISCUSSION

We report initial XBB.1.5 vaccine effectiveness of 64% against severe outcomes, relative to prior non-XBB.1.5 vaccines among community-dwelling adults, when the XBB sublineage predominated. Entering the JN/KP-predominant period, rVE was reduced to 57% and quickly declined as time elapsed. No significant benefit was observed beyond 6 months of vaccination. Our results are largely consistent with early reports describing VE of 52%−67% against severe illness during fall/winter 2023 [5-11]. However, we found brief durability of protection, underscoring the potential need for vaccination more than once per year to prevent COVID-19-associated severe outcomes [12]. Given the rapid evolution of the Omicron variant, by the time the XBB.1.5 vaccine became available, the immune-evasive JN/KP sublineages had started to emerge and quickly replaced the XBB sublineage within a few months. Although some degree of cross-protection may exist initially [8,9], it waned within half a year. The new KP.2-adapted vaccine did not become available until fall 2024 (i.e., almost one year after the XBB.1.5 vaccine). Since seasonality of COVID-19 has not been established, our results indicate the urgency for swift composition updates and deployment of the latest adapted vaccine, especially for high-risk populations regardless of time of the year or vaccination history.

Notably, our analyses indicate that rVE of an updated vaccine can be influenced by the timing and type of vaccines received by the reference group. We observed higher estimates when the last vaccine dose was received more remotely (i.e., ≥12 months ago) by the reference group; and likewise, when it was the original monovalent rather than the omicron-containing bivalent vaccine (although the more recent timing of the latter could have contributed). We included estimates of absolute VE because it better reflects the full benefit of vaccination, however for highly vaccinated populations this measure may not inform vaccination strategies.

Limitations of our study include restricted access to PCR testing during the study period, unavailability of data on rapid antigen tests and individual-level whole genome sequencing results, inability to separate the impact of virus evolution and time since vaccination, and the possibility of residual confounding.

Continual monitoring of the effectiveness of updated COVID-19 vaccines as both SARS-CoV-2 and population immunity evolve is critical. More durable and antigenically conserved COVID-19 vaccines are necessary.

## Data Availability

All data produced in the present work are contained in the manuscript

## Acknowledgements and Disclaimers

We would like to acknowledge Public Health Ontario for access to vaccination data from COVaxON, case-level data from the Public Health Case and Contact Management Solution (CCM) and COVID-19 laboratory data, aggregate whole genome sequencing data, as well as assistance with data interpretation. We also thank the staff of Ontario’s public health units who are responsible for COVID-19 case and contact management and data collection within CCM. We thank IQVIA Solutions Canada Inc. for use of their Drug Information File. The authors are grateful to the Ontario residents without whom this research would be impossible.

This document used data adapted from the Statistics Canada Postal Code^OM^ Conversion File, which is based on data licensed from Canada Post Corporation, and/or data adapted from the Ontario Ministry of Health Postal Code Conversion File, which contains data copied under license from ^©^Canada Post Corporation and Statistics Canada. Parts of this material are based on data and/or information compiled and provided by: MOH, Ontario Health, the Canadian Institute for Health Information, Statistics Canada, and IQVIA Solutions Canada Inc. The analyses, conclusions, opinions and statements expressed herein are solely those of the authors and do not reflect those of the funding or data sources; no endorsement is intended or should be inferred. Adapted from Statistics Canada, Canadian Census 2016. This does not constitute an endorsement by Statistics Canada of this product. The use of ICES data in this study is authorized under section 45 of Ontario’s Personal Health Information Protection Act and does not require review by a research ethics board.

## Appendix

**Figure S1.**
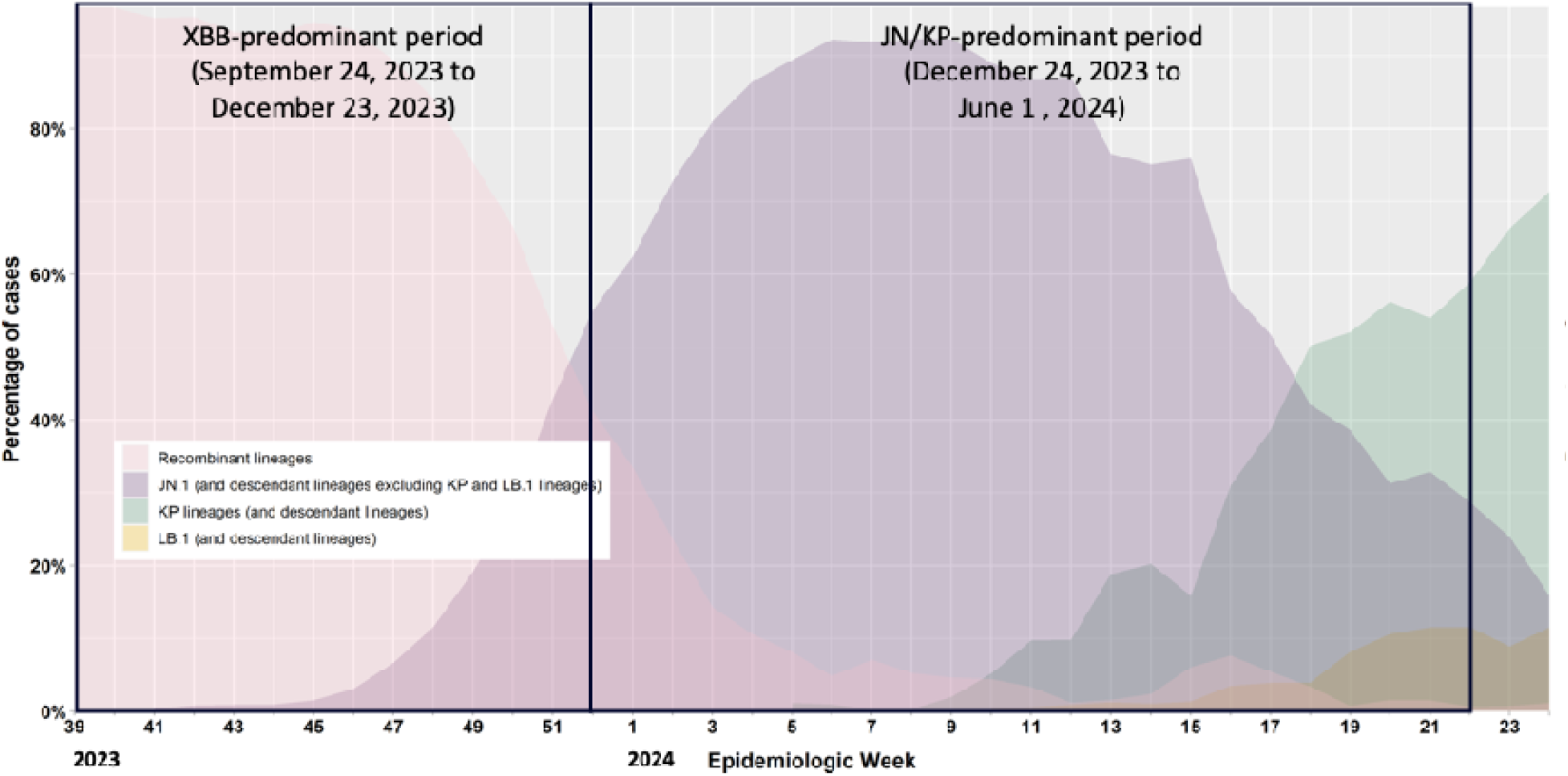
Percentage of COVID-19 cases by the most prevalent Omicron sublineages and week in Ontario, 24 September 2023 to 1 June 2024

**Table S1.** Descriptive characteristics of community-dwelling adults aged ≥50 years tested for SARS-CoV-2 between 24 September 2023 and 1 June 2024 in Ontario, Canada, comparing cases with Omicron-associated severe outcomes and test-negative controls.

|  | XBB-predominant period<br>(24 Sep, 2023 – 23 Dec, 2023) |  |  | JN/KP-predominant period<br>(24 Dec, 2023 – 1 Jun 2024) |  |  |
| --- | --- | --- | --- | --- | --- | --- |
|  | Omicron cases,<br>n (%)<sup>a</sup> | Test-negative controls,<br>n (%)<sup>a</sup> | SD<sup>b</sup> | Omicron cases,<br>n (%)<sup>a</sup> | Test-negative controls,<br>n (%)<sup>a</sup> | SD<sup>b</sup> |
|  | N=3,093 | N=8,194 |  | N=1,802 | N=15,029 |  |
| Mean age (standard deviation), years | 80.00 (10.75) | 72.43 (12.84) | 0.64 | 79.26 (10.75) | 73.47 (12.58) | 0.50 |
| Age group, years |  |  |  |  |  |  |
| 50-64 | 318 (10.3%) | 2,665 (32.5%) | 0.56 | 201 (11.2%) | 4,252 (28.3%) | 0.44 |
| 65-74 | 558 (18.0%) | 1,801 (22.0%) | 0.10 | 340 (18.9%) | 3,486 (23.2%) | 0.11 |
| 75-84 | 1,012 (32.7%) | 1,941 (23.7%) | 0.20 | 598 (33.2%) | 3,760 (25.0%) | 0.18 |
| $\geq 85$ | 1,205 (39.0%) | 1,787 (21.8%) | 0.38 | 663 (36.8%) | 3,531 (23.5%) | 0.29 |
| Male sex | 1,643 (53.1%) | 3,454 (42.2%) | 0.22 | 995 (55.2%) | 6,547 (43.6%) | 0.24 |
| Public health unit region |  |  |  |  |  |  |
| Central East | 109 (3.5%) | 201 (2.5%) | 0.06 | 58 (3.2%) | 346 (2.3%) | 0.06 |
| Central West | 869 (28.1%) | 1,659 (20.2%) | 0.18 | 474 (26.3%) | 2,544 (16.9%) | 0.23 |
| Durham | 82 (2.7%) | 69 (0.8%) | 0.14 | 32 (1.8%) | 96 (0.6%) | 0.10 |
| Eastern | 496 (16.0%) | 409 (5.0%) | 0.37 | 241 (13.4%) | 763 (5.1%) | 0.29 |
| North | 269 (8.7%) | 2,261 (27.6%) | 0.51 | 238 (13.2%) | 4,414 (29.4%) | 0.40 |
| Ottawa | 45 (1.5%) | 503 (6.1%) | 0.25 | 26 (1.4%) | 685 (4.6%) | 0.18 |
| Peel | 37 (1.2%) | 858 (10.5%) | 0.40 | 27 (1.5%) | 1,048 (7.0%) | 0.27 |
| South West | 266 (8.6%) | 1,460 (17.8%) | 0.28 | 139 (7.7%) | 4,044 (26.9%) | 0.52 |
| Toronto | 642 (20.8%) | 506 (6.2%) | 0.44 | 390 (21.6%) | 661 (4.4%) | 0.53 |
| York | 224 (7.2%) | 120 (1.5%) | 0.29 | 140 (7.8%) | 164 (1.1%) | 0.33 |
| Missing | 54 (1.7%) | 148 (1.8%) | 0.01 | 37 (2.1%) | 264 (1.8%) | 0.02 |
| Household income quintile |  |  |  |  |  |  |
| 1 (lowest) | 912 (29.5%) | 2,123 (25.9%) | 0.08 | 522 (29.0%) | 4,143 (27.6%) | 0.03 |
|  | Omicron cases,<br>n (%)<sup>a</sup> | Test-negative controls,<br>n (%)<sup>a</sup> | SD<sup>b</sup> | Omicron cases,<br>n (%)<sup>a</sup> | Test-negative controls,<br>n (%)<sup>a</sup> | SD<sup>b</sup> |
| 2 | 706 (22.8%) | 1,773 (21.6%) | 0.03 | 388 (21.5%) | 3,259 (21.7%) | 0.00 |
| 3 | 584 (18.9%) | 1,576 (19.2%) | 0.01 | 353 (19.6%) | 2,812 (18.7%) | 0.02 |
| 4 | 456 (14.7%) | 1,397 (17.0%) | 0.06 | 284 (15.8%) | 2,515 (16.7%) | 0.03 |
| 5 (highest) | 427 (13.8%) | 1,271 (15.5%) | 0.05 | 248 (13.8%) | 2,222 (14.8%) | 0.03 |
| Missing | 8 (0.3%) | 54 (0.7%) | 0.06 | 7 (0.4%) | 78 (0.5%) | 0.02 |
| Essential workers quintile |  |  |  |  |  |  |
| 1 (0–32.5) | 407 (13.2%) | 1,081 (13.2%) | 0.00 | 247 (13.7%) | 1,783 (11.9%) | 0.06 |
| 2 (32.5–42.3) | 610 (19.7%) | 1,655 (20.2%) | 0.01 | 318 (17.6%) | 2,677 (17.8%) | 0.00 |
| 3 (42.3–49.8) | 625 (20.2%) | 1,700 (20.7%) | 0.01 | 356 (19.8%) | 3,164 (21.1%) | 0.03 |
| 4 (50.0–57.5) | 648 (21.0%) | 1,649 (20.1%) | 0.02 | 384 (21.3%) | 3,194 (21.3%) | 0.00 |
| 5 (57.5–100) | 650 (21.0%) | 1,649 (20.1%) | 0.02 | 399 (22.1%) | 3,480 (23.2%) | 0.02 |
| Missing | 153 (4.9%) | 460 (5.6%) | 0.03 | 98 (5.4%) | 731 (4.9%) | 0.03 |
| Persons per dwelling quintile |  |  |  |  |  |  |
| 1 (0–2.1) | 920 (29.7%) | 2,498 (30.5%) | 0.02 | 504 (28.0%) | 4,811 (32.0%) | 0.09 |
| 2 (2.2–2.4) | 712 (23.0%) | 1,825 (22.3%) | 0.02 | 406 (22.5%) | 3,582 (23.8%) | 0.03 |
| 3 (2.5–2.6) | 414 (13.4%) | 1,123 (13.7%) | 0.01 | 245 (13.6%) | 2,035 (13.5%) | 0.00 |
| 4 (2.7–3.0) | 592 (19.1%) | 1,359 (16.6%) | 0.07 | 354 (19.6%) | 2,459 (16.4%) | 0.09 |
| 5 (3.1–5.7) | 304 (9.8%) | 934 (11.4%) | 0.05 | 196 (10.9%) | 1,416 (9.4%) | 0.05 |
| Missing | 151 (4.9%) | 455 (5.6%) | 0.03 | 97 (5.4%) | 726 (4.8%) | 0.03 |
| Self-identified visible minority quintile |  |  |  |  |  |  |
| 1 (0.0–2.2) | 689 (22.3%) | 2,139 (26.1%) | 0.09 | 446 (24.8%) | 4,200 (27.9%) | 0.07 |
| 2 (2.2–7.5) | 702 (22.7%) | 1,898 (23.2%) | 0.01 | 383 (21.3%) | 3,780 (25.2%) | 0.09 |
| 3 (7.5–18.7) | 584 (18.9%) | 1,493 (18.2%) | 0.02 | 321 (17.8%) | 2,832 (18.8%) | 0.03 |
| 4 (18.7–43.5) | 492 (15.9%) | 1,383 (16.9%) | 0.03 | 287 (15.9%) | 2,321 (15.4%) | 0.01 |
|  | Omicron cases,<br>n (%)<sup>a</sup> | Test-negative controls,<br>n (%)<sup>a</sup> | SD<sup>b</sup> | Omicron cases,<br>n (%)<sup>a</sup> | Test-negative controls,<br>n (%)<sup>a</sup> | SD<sup>b</sup> |
| 5 (43.5–100) | 475 (15.4%) | 822 (10.0%) | 0.16 | 268 (14.9%) | 1,168 (7.8%) | 0.23 |
| Missing | 151 (4.9%) | 459 (5.6%) | 0.03 | 97 (5.4%) | 728 (4.8%) | 0.02 |
| Receipt of 2022-2023 and/or 2023-2024 influenza vaccination | 1,420 (45.9%) | 3,313 (40.4%) | 0.11 | 802 (44.5%) | 6,247 (41.6%) | 0.06 |
| Any comorbidity (other than immunocompromised) | 2,931 (94.8%) | 7,044 (86.0%) | 0.30 | 1,686 (93.6%) | 13,140 (87.4%) | 0.21 |
| Receipt of home care services |  |  |  |  |  |  |
| None | 2,693 (87.1%) | 7,476 (91.2%) | 0.13 | 1,562 (86.7%) | 13,410 (89.2%) | 0.08 |
| Short stay | 118 (3.8%) | 322 (3.9%) | 0.01 | 71 (3.9%) | 777 (5.2%) | 0.06 |
| Long stay | 275 (8.9%) | 379 (4.6%) | 0.17 | 162 (9.0%) | 772 (5.1%) | 0.15 |
| Palliative | 7 (0.2%) | 17 (0.2%) | 0.00 | 7 (0.4%) | 70 (0.5%) | 0.01 |
| Positive SARS-CoV-2 PCR test, >60 days prior) | 385 (12.4%) | 2,157 (26.3%) | 0.36 | 247 (13.7%) | 3,248 (21.6%) | 0.21 |
| Most recent vaccine dose |  |  |  |  |  |  |
| Unvaccinated | 263 (8.5%) | 524 (6.4%) | 0.08 | 170 (9.4%) | 1,103 (7.3%) | 0.08 |
| Original monovalent | 1,780 (57.5%) | 4,946 (60.4%) | 0.06 | 1,320 (73.3%) | 11,674 (77.7%) | 0.10 |
| Omicron-containing bivalent | 1,050 (33.9%) | 2,724 (33.2%) | 0.02 | 312 (17.3%) | 2,252 (15.0%) | 0.06 |
| Mean time since last dose (standard deviation), days | 465.4 (247.5) | 422.7 (270.5) | 0.17 | 493.6 (306.5) | 466.0 (330.2) | 0.09 |

<sup>a</sup> Proportion reported, unless stated otherwise. <sup>b</sup> SD = standardized difference. Standardized differences of >0.10 are considered clinically relevant. Cases with Omicron-associated severe outcomes were compared with test-negative controls in each Omicron sublineage-predominant period.

**Table S2.**
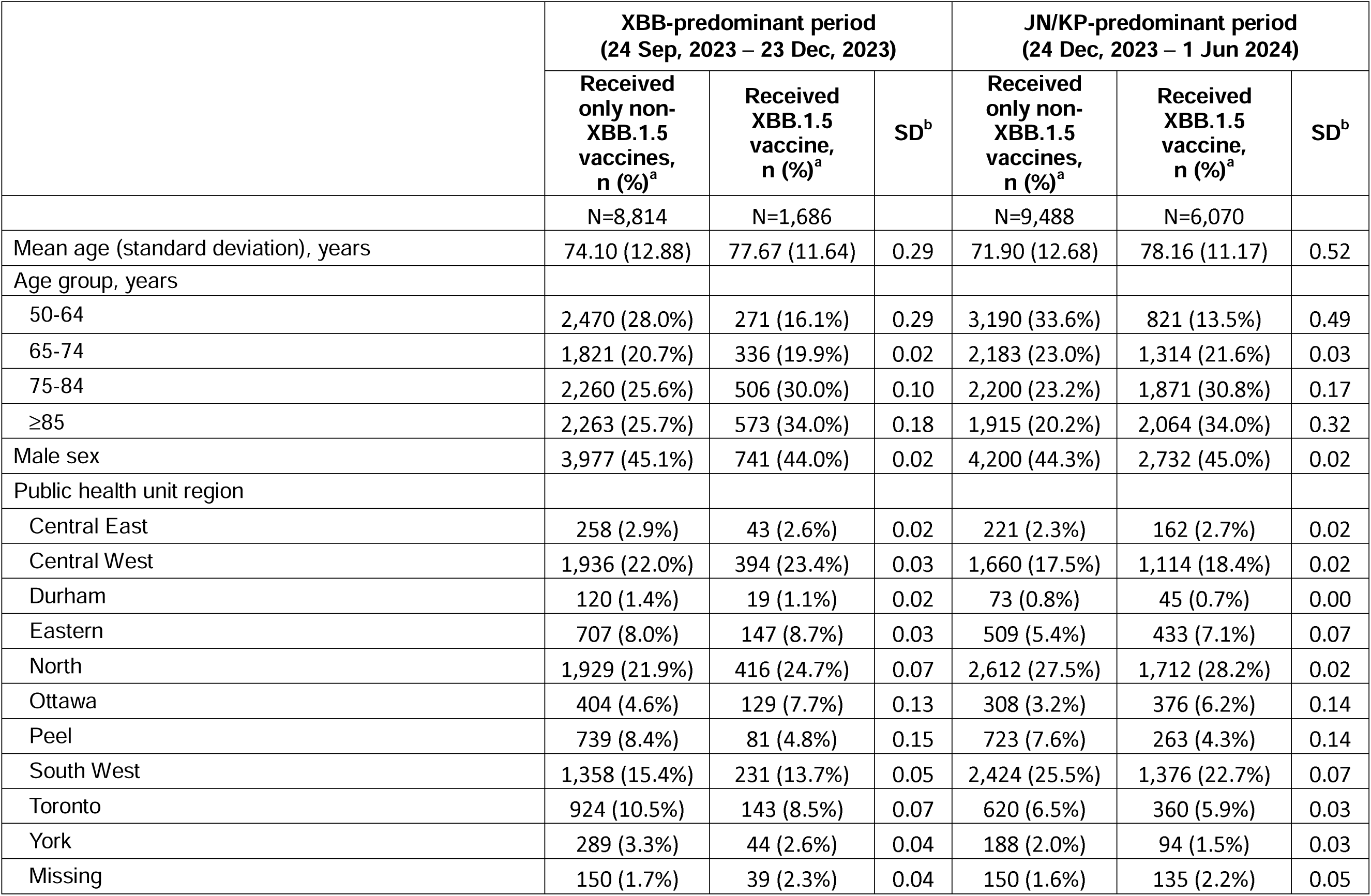
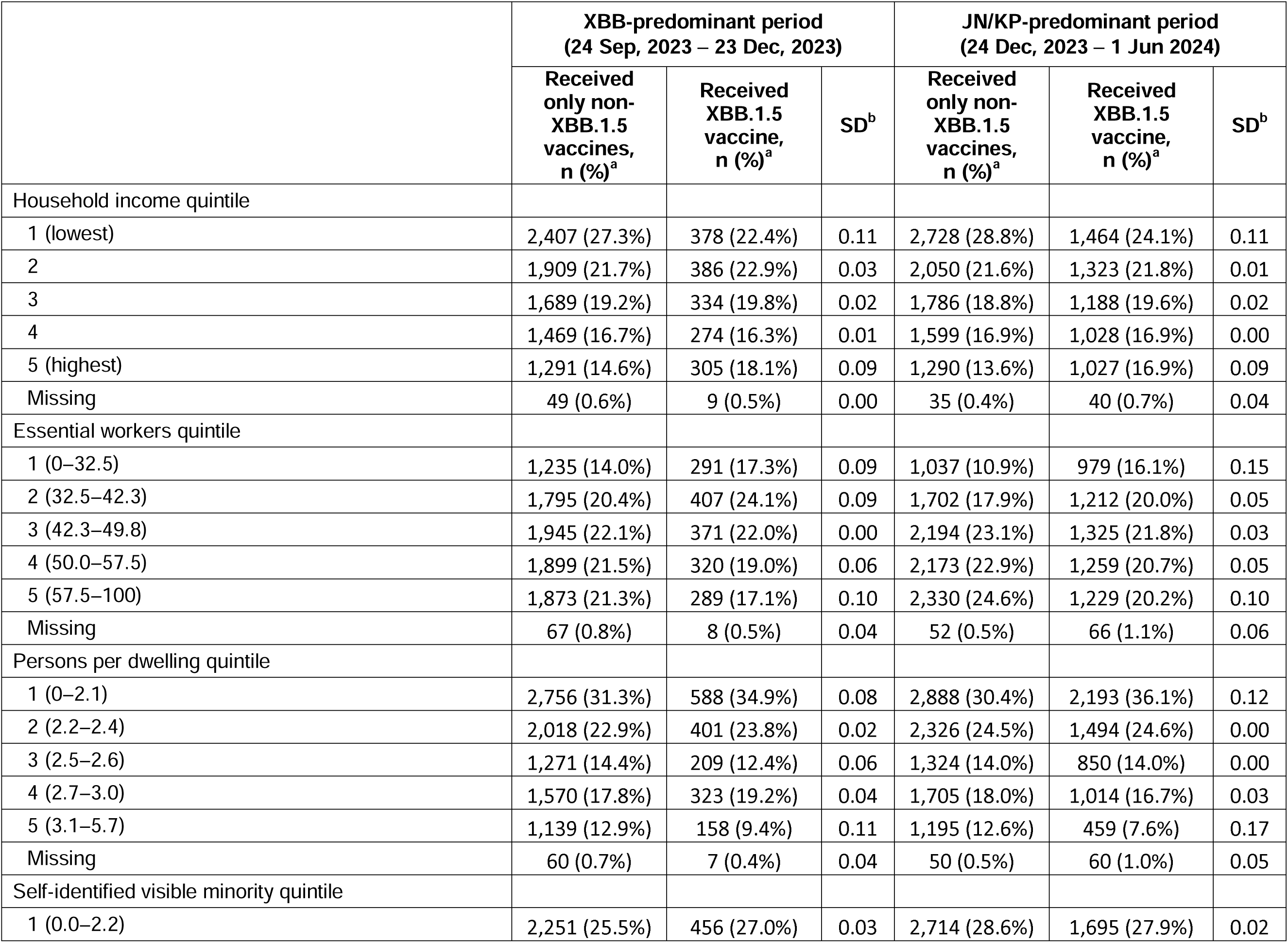
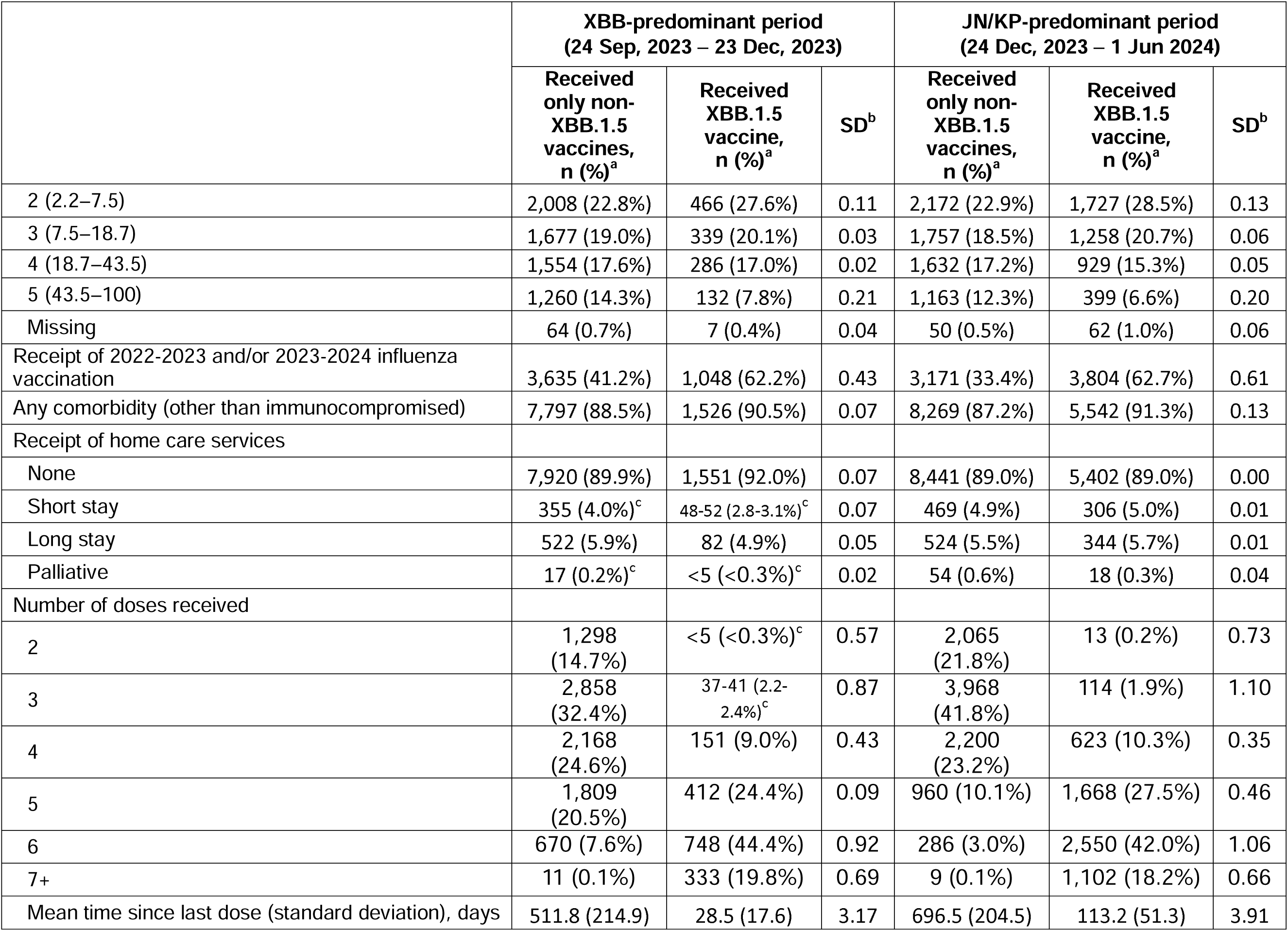
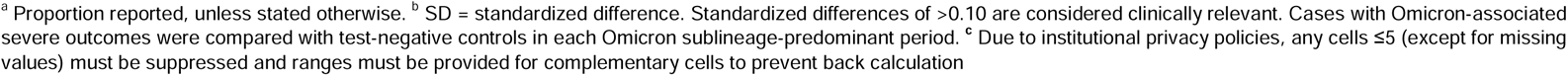
Descriptive characteristics of community-dwelling adults aged ≥50 years tested for SARS-CoV-2 between 24 September 2023 and 1 June 2024 in Ontario, Canada, comparing subjects who received XBB.1.5 vaccines and subjects who received only ≥2 doses of non-XBB.1.5 vaccines.

|  | XBB-predominant period<br>(24 Sep, 2023 – 23 Dec, 2023) |  |  | JN/KP-predominant period<br>(24 Dec, 2023 – 1 Jun 2024) |  |  |
| --- | --- | --- | --- | --- | --- | --- |
|  | Received<br>only non-XBB.1.5<br>vaccines,<br>n (%)<sup>a</sup> | Received<br>XBB.1.5<br>vaccine,<br>n (%)<sup>a</sup> | SD<sup>b</sup> | Received<br>only non-XBB.1.5<br>vaccines,<br>n (%)<sup>a</sup> | Received<br>XBB.1.5<br>vaccine,<br>n (%)<sup>a</sup> | SD<sup>b</sup> |
|  | N=8,814 | N=1,686 |  | N=9,488 | N=6,070 |  |
| Mean age (standard deviation), years | 74.10 (12.88) | 77.67 (11.64) | 0.29 | 71.90 (12.68) | 78.16 (11.17) | 0.52 |
| Age group, years |  |  |  |  |  |  |
| 50-64 | 2,470 (28.0%) | 271 (16.1%) | 0.29 | 3,190 (33.6%) | 821 (13.5%) | 0.49 |
| 65-74 | 1,821 (20.7%) | 336 (19.9%) | 0.02 | 2,183 (23.0%) | 1,314 (21.6%) | 0.03 |
| 75-84 | 2,260 (25.6%) | 506 (30.0%) | 0.10 | 2,200 (23.2%) | 1,871 (30.8%) | 0.17 |
| ≥85 | 2,263 (25.7%) | 573 (34.0%) | 0.18 | 1,915 (20.2%) | 2,064 (34.0%) | 0.32 |
| Male sex | 3,977 (45.1%) | 741 (44.0%) | 0.02 | 4,200 (44.3%) | 2,732 (45.0%) | 0.02 |
| Public health unit region |  |  |  |  |  |  |
| Central East | 258 (2.9%) | 43 (2.6%) | 0.02 | 221 (2.3%) | 162 (2.7%) | 0.02 |
| Central West | 1,936 (22.0%) | 394 (23.4%) | 0.03 | 1,660 (17.5%) | 1,114 (18.4%) | 0.02 |
| Durham | 120 (1.4%) | 19 (1.1%) | 0.02 | 73 (0.8%) | 45 (0.7%) | 0.00 |
| Eastern | 707 (8.0%) | 147 (8.7%) | 0.03 | 509 (5.4%) | 433 (7.1%) | 0.07 |
| North | 1,929 (21.9%) | 416 (24.7%) | 0.07 | 2,612 (27.5%) | 1,712 (28.2%) | 0.02 |
| Ottawa | 404 (4.6%) | 129 (7.7%) | 0.13 | 308 (3.2%) | 376 (6.2%) | 0.14 |
| Peel | 739 (8.4%) | 81 (4.8%) | 0.15 | 723 (7.6%) | 263 (4.3%) | 0.14 |
| South West | 1,358 (15.4%) | 231 (13.7%) | 0.05 | 2,424 (25.5%) | 1,376 (22.7%) | 0.07 |
| Toronto | 924 (10.5%) | 143 (8.5%) | 0.07 | 620 (6.5%) | 360 (5.9%) | 0.03 |
| York | 289 (3.3%) | 44 (2.6%) | 0.04 | 188 (2.0%) | 94 (1.5%) | 0.03 |
| Missing | 150 (1.7%) | 39 (2.3%) | 0.04 | 150 (1.6%) | 135 (2.2%) | 0.05 |
|  | Received only non-XBB.1.5 vaccines, n (%)<sup>a</sup> | Received XBB.1.5 vaccine, n (%)<sup>a</sup> | SD<sup>b</sup> | Received only non-XBB.1.5 vaccines, n (%)<sup>a</sup> | Received XBB.1.5 vaccine, n (%)<sup>a</sup> | SD<sup>b</sup> |
| Household income quintile |  |  |  |  |  |  |
| 1 (lowest) | 2,407 (27.3%) | 378 (22.4%) | 0.11 | 2,728 (28.8%) | 1,464 (24.1%) | 0.11 |
| 2 | 1,909 (21.7%) | 386 (22.9%) | 0.03 | 2,050 (21.6%) | 1,323 (21.8%) | 0.01 |
| 3 | 1,689 (19.2%) | 334 (19.8%) | 0.02 | 1,786 (18.8%) | 1,188 (19.6%) | 0.02 |
| 4 | 1,469 (16.7%) | 274 (16.3%) | 0.01 | 1,599 (16.9%) | 1,028 (16.9%) | 0.00 |
| 5 (highest) | 1,291 (14.6%) | 305 (18.1%) | 0.09 | 1,290 (13.6%) | 1,027 (16.9%) | 0.09 |
| Missing | 49 (0.6%) | 9 (0.5%) | 0.00 | 35 (0.4%) | 40 (0.7%) | 0.04 |
| Essential workers quintile |  |  |  |  |  |  |
| 1 (0–32.5) | 1,235 (14.0%) | 291 (17.3%) | 0.09 | 1,037 (10.9%) | 979 (16.1%) | 0.15 |
| 2 (32.5–42.3) | 1,795 (20.4%) | 407 (24.1%) | 0.09 | 1,702 (17.9%) | 1,212 (20.0%) | 0.05 |
| 3 (42.3–49.8) | 1,945 (22.1%) | 371 (22.0%) | 0.00 | 2,194 (23.1%) | 1,325 (21.8%) | 0.03 |
| 4 (50.0–57.5) | 1,899 (21.5%) | 320 (19.0%) | 0.06 | 2,173 (22.9%) | 1,259 (20.7%) | 0.05 |
| 5 (57.5–100) | 1,873 (21.3%) | 289 (17.1%) | 0.10 | 2,330 (24.6%) | 1,229 (20.2%) | 0.10 |
| Missing | 67 (0.8%) | 8 (0.5%) | 0.04 | 52 (0.5%) | 66 (1.1%) | 0.06 |
| Persons per dwelling quintile |  |  |  |  |  |  |
| 1 (0–2.1) | 2,756 (31.3%) | 588 (34.9%) | 0.08 | 2,888 (30.4%) | 2,193 (36.1%) | 0.12 |
| 2 (2.2–2.4) | 2,018 (22.9%) | 401 (23.8%) | 0.02 | 2,326 (24.5%) | 1,494 (24.6%) | 0.00 |
| 3 (2.5–2.6) | 1,271 (14.4%) | 209 (12.4%) | 0.06 | 1,324 (14.0%) | 850 (14.0%) | 0.00 |
| 4 (2.7–3.0) | 1,570 (17.8%) | 323 (19.2%) | 0.04 | 1,705 (18.0%) | 1,014 (16.7%) | 0.03 |
| 5 (3.1–5.7) | 1,139 (12.9%) | 158 (9.4%) | 0.11 | 1,195 (12.6%) | 459 (7.6%) | 0.17 |
| Missing | 60 (0.7%) | 7 (0.4%) | 0.04 | 50 (0.5%) | 60 (1.0%) | 0.05 |
| Self-identified visible minority quintile |  |  |  |  |  |  |
| 1 (0.0–2.2) | 2,251 (25.5%) | 456 (27.0%) | 0.03 | 2,714 (28.6%) | 1,695 (27.9%) | 0.02 |
|  | Received only non-XBB.1.5 vaccines, n (%)<sup>a</sup> | Received XBB.1.5 vaccine, n (%)<sup>a</sup> | SD<sup>b</sup> | Received only non-XBB.1.5 vaccines, n (%)<sup>a</sup> | Received XBB.1.5 vaccine, n (%)<sup>a</sup> | SD<sup>b</sup> |
| 2 (2.2–7.5) | 2,008 (22.8%) | 466 (27.6%) | 0.11 | 2,172 (22.9%) | 1,727 (28.5%) | 0.13 |
| 3 (7.5–18.7) | 1,677 (19.0%) | 339 (20.1%) | 0.03 | 1,757 (18.5%) | 1,258 (20.7%) | 0.06 |
| 4 (18.7–43.5) | 1,554 (17.6%) | 286 (17.0%) | 0.02 | 1,632 (17.2%) | 929 (15.3%) | 0.05 |
| 5 (43.5–100) | 1,260 (14.3%) | 132 (7.8%) | 0.21 | 1,163 (12.3%) | 399 (6.6%) | 0.20 |
| Missing | 64 (0.7%) | 7 (0.4%) | 0.04 | 50 (0.5%) | 62 (1.0%) | 0.06 |
| Receipt of 2022-2023 and/or 2023-2024 influenza vaccination | 3,635 (41.2%) | 1,048 (62.2%) | 0.43 | 3,171 (33.4%) | 3,804 (62.7%) | 0.61 |
| Any comorbidity (other than immunocompromised) | 7,797 (88.5%) | 1,526 (90.5%) | 0.07 | 8,269 (87.2%) | 5,542 (91.3%) | 0.13 |
| Receipt of home care services |  |  |  |  |  |  |
| None | 7,920 (89.9%) | 1,551 (92.0%) | 0.07 | 8,441 (89.0%) | 5,402 (89.0%) | 0.00 |
| Short stay | 355 (4.0%)<sup>c</sup> | 48-52 (2.8-3.1%)<sup>c</sup> | 0.07 | 469 (4.9%) | 306 (5.0%) | 0.01 |
| Long stay | 522 (5.9%) | 82 (4.9%) | 0.05 | 524 (5.5%) | 344 (5.7%) | 0.01 |
| Palliative | 17 (0.2%)<sup>c</sup> | <5 (<0.3%)<sup>c</sup> | 0.02 | 54 (0.6%) | 18 (0.3%) | 0.04 |
| Number of doses received |  |  |  |  |  |  |
| 2 | 1,298 (14.7%) | <5 (<0.3%)<sup>c</sup> | 0.57 | 2,065 (21.8%) | 13 (0.2%) | 0.73 |
| 3 | 2,858 (32.4%) | 37-41 (2.2-2.4%)<sup>c</sup> | 0.87 | 3,968 (41.8%) | 114 (1.9%) | 1.10 |
| 4 | 2,168 (24.6%) | 151 (9.0%) | 0.43 | 2,200 (23.2%) | 623 (10.3%) | 0.35 |
| 5 | 1,809 (20.5%) | 412 (24.4%) | 0.09 | 960 (10.1%) | 1,668 (27.5%) | 0.46 |
| 6 | 670 (7.6%) | 748 (44.4%) | 0.92 | 286 (3.0%) | 2,550 (42.0%) | 1.06 |
| 7+ | 11 (0.1%) | 333 (19.8%) | 0.69 | 9 (0.1%) | 1,102 (18.2%) | 0.66 |
| Mean time since last dose (standard deviation), days | 511.8 (214.9) | 28.5 (17.6) | 3.17 | 696.5 (204.5) | 113.2 (51.3) | 3.91 |

<sup>a</sup> Proportion reported, unless stated otherwise. <sup>b</sup> SD = standardized difference. Standardized differences of >0.10 are considered clinically relevant. Cases with Omicron-associated severe outcomes were compared with test-negative controls in each Omicron sublineage-predominant period. <sup>c</sup> Due to institutional privacy policies, any cells $\leq 5$ (except for missing values) must be suppressed and ranges must be provided for complementary cells to prevent back calculation

**Figure S2.**
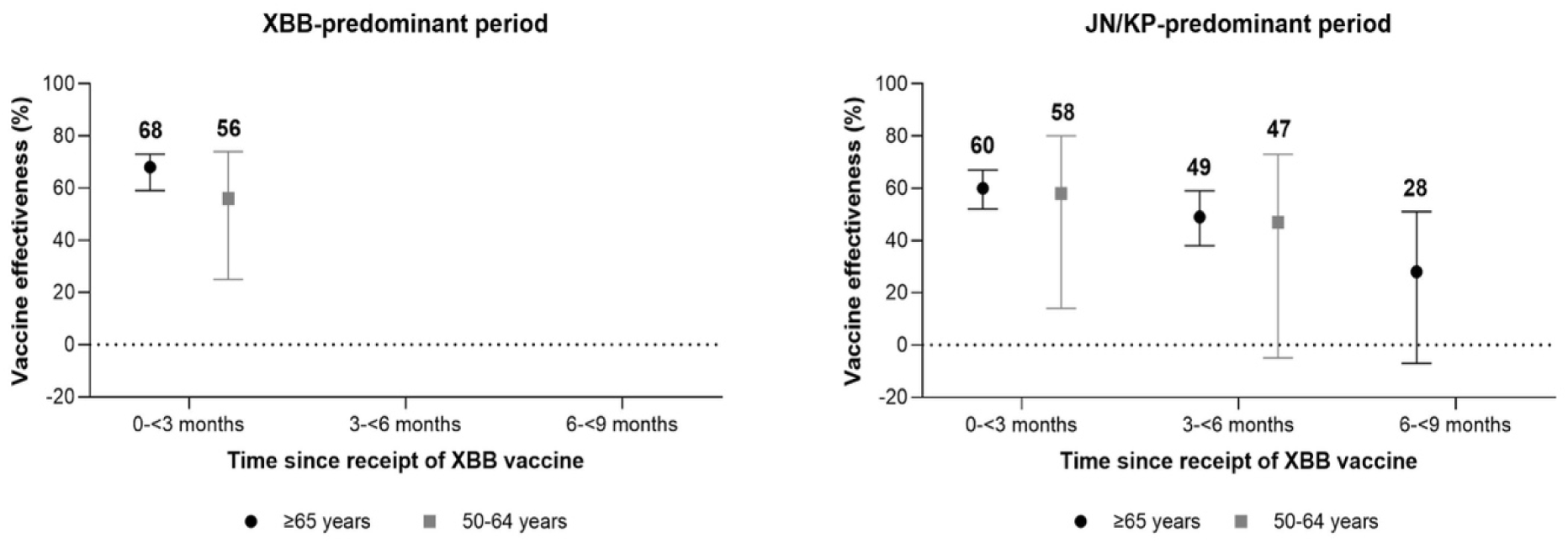
Relative vaccine effectiveness of mRNA XBB.1.5 vaccination over time, compared to subjects who received only non-XBB.1.5 vaccines at least 6 months prior, during periods of XBB and JN/KP sublineage predominance, stratified by age group.

**Figure S3.**
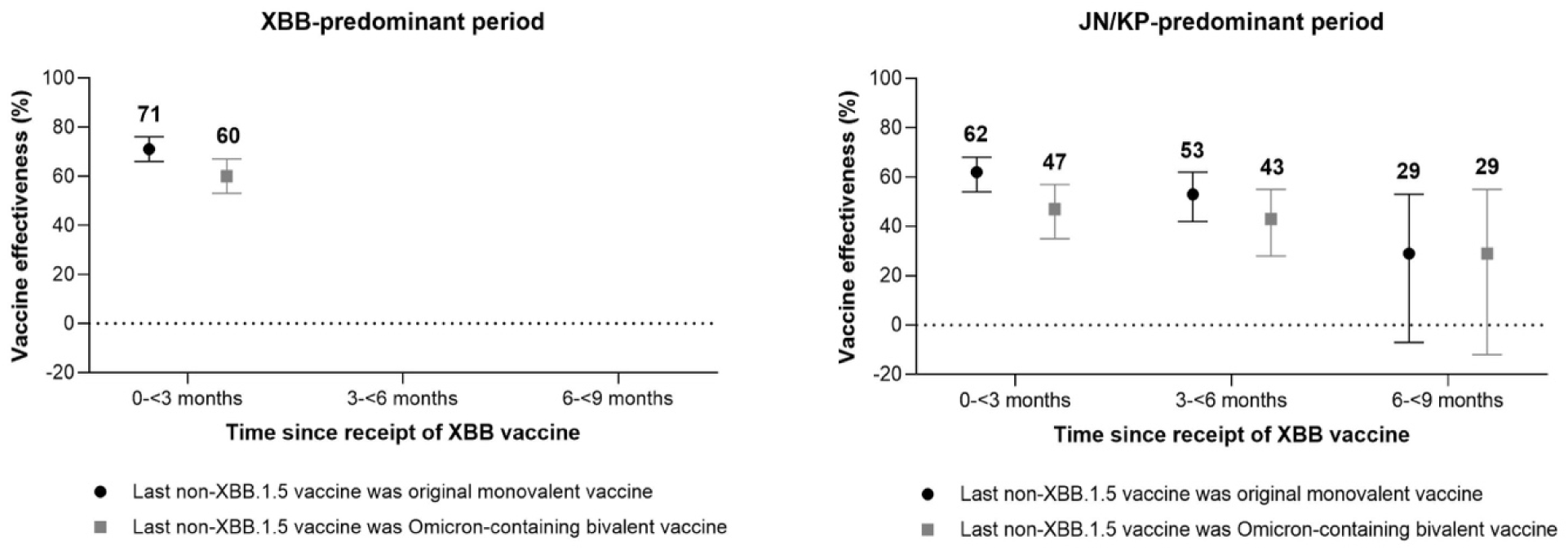
Relative vaccine effectiveness of mRNA XBB.1.5 vaccination over time, compared to subjects who received only non-XBB.1.5 vaccines at least 6 months prior, during periods of XBB and JN/KP sublineage predominance, stratified by the type of non-XBB.1.5 vaccine last received.

**Figure S4.**
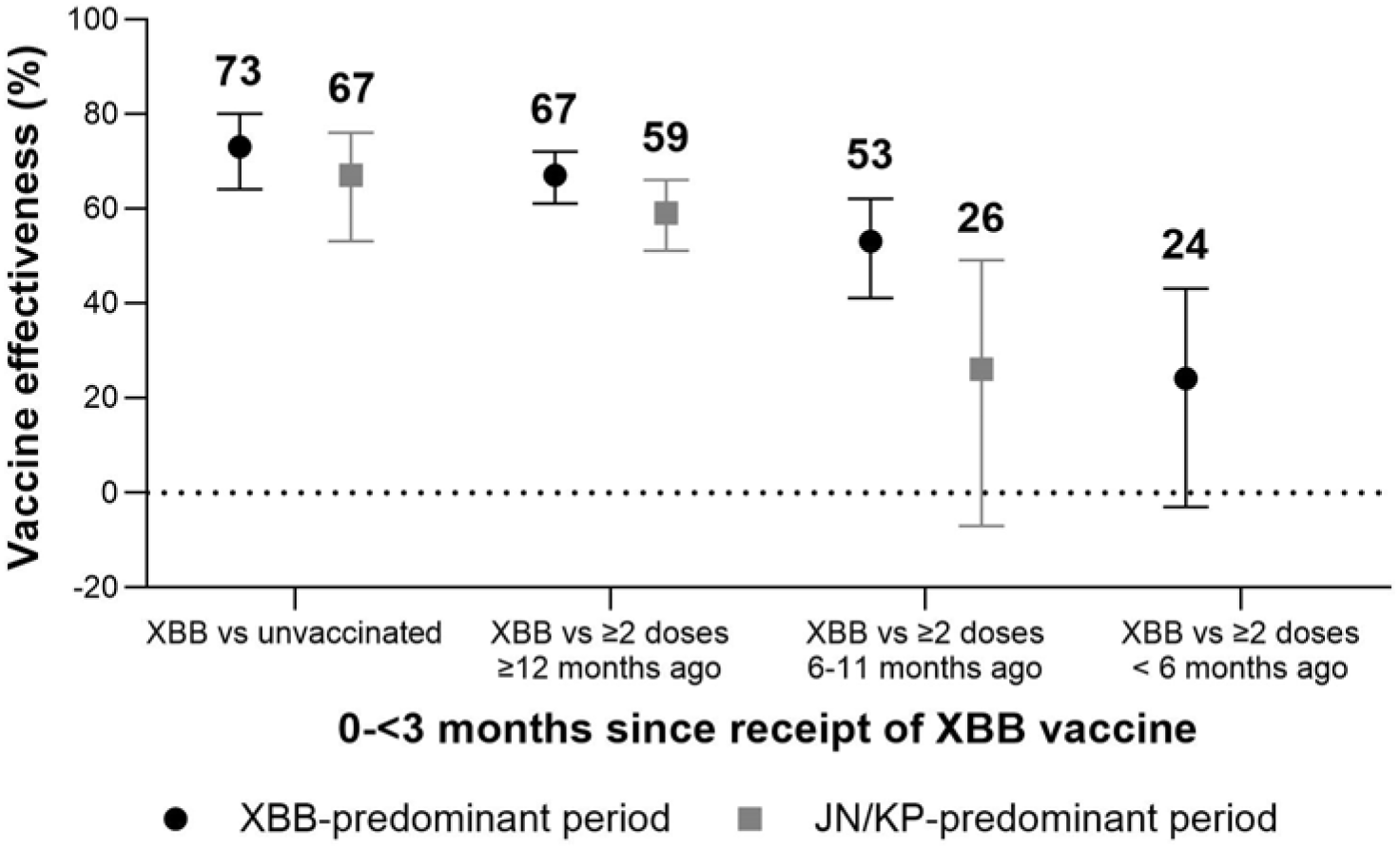
Effectiveness of mRNA XBB.1.5 vaccination after 0−<3 months, compared to various reference groups

## References

1. Lee N, Nguyen L, Austin PC, Brown KA, Grewal R, Buchan SA, Nasreen S, Gubbay J, Schwartz KL, Tadrous M, Wilson K, Wilson SE, Kwong JC. Protection Conferred by COVID-19 Vaccination, Prior SARS-CoV-2 Infection, or Hybrid Immunity Against Omicron-Associated Severe Outcomes Among Community-Dwelling Adults. Clin Infect Dis. 2024;78(5):1372–1382.

2. Grewal R, Buchan SA, Nguyen L, Nasreen S, Austin PC, Brown KA, Gubbay J, Lee N, Schwartz KL, Tadrous M, Wilson K, Wilson SE, Kwong JC. Effectiveness of mRNA COVID-19 Monovalent and Bivalent Vaccine Booster Doses Against Omicron Severe Outcomes Among Adults Aged 50 Years in Ontario, Canada: A Canadian Immunization Research Network Study. J Infect Dis. 2024;229(2):394-397.

3. National Advisory Committee on Immunization (NACI). An Advisory Committee Statement (ACS). Addendum to the guidance on the use of COVID-19 vaccines in the fall of 2023. https://www.canada.ca/en/public-health/services/publications/vaccines-immunization/national-advisory-committee-immunization-addendum-guidance-use-covid-19-vaccines-fall-2023.html

4. Government of Ontario. COVID-19 Vaccine Guidance Version 11.1 – April 8, 2024. https://www.ontario.ca/files/2024-04/moh-covid-19-vaccine-guidance-en-2024-04-08.pdf

5. Tartof SY, Slezak JM, Frankland TB, Puzniak L, Hong V, Ackerson BK, Stern JA, Zamparo J, Simmons S, Jodar L, McLaughlin JM. Estimated Effectiveness of the BNT162b2 XBB Vaccine Against COVID-19. JAMA Intern Med. 2024;184(8):932–940.

6. Kirsebom FCM, Stowe J, Lopez Bernal J, Allen A, Andrews N. Effectiveness of autumn 2023 COVID-19 vaccination and residual protection of prior doses against hospitalisation in England, estimated using a test-negative case-control study. J Infect. 2024;89(1):106177.

7. Antunes L, Mazagatos C, Martínez-Baz I, Naesens R, Borg ML, Petrović G, et al.; European Hospital Vaccine Effectiveness Group. Early COVID-19 XBB.1.5 Vaccine Effectiveness Against Hospitalisation Among Adults Targeted for Vaccination, VEBIS Hospital Network, Europe, October 2023-January 2024. Influenza Other Respir Viruses. 2024;18(8):e13360.

8. Ma KC, Surie D, Lauring AS, Martin ET, Leis AM, Papalambros L, et al. Effectiveness of Updated 2023-2024 (Monovalent XBB.1.5) COVID-19 Vaccination Against SARS-CoV-2 Omicron XBB and BA.2.86/JN.1 Lineage Hospitalization and a Comparison of Clinical Severity-IVY Network, 26 Hospitals, October 18, 2023-March 9, 2024. Clin Infect Dis. 2024:ciae405.

9. Lin DY, Du Y, Xu Y, Paritala S, Donahue M, Maloney P. Durability of XBB.1.5 Vaccines against Omicron Subvariants. N Engl J Med. 2024;390(22):2124–2127.

10. Monge S, Humphreys J, Nicolay N, Braeye T, Van Evercooren I, Holm Hansen C, et al.; VEBIS-EHR Working Group. Effectiveness of XBB.1.5 Monovalent COVID-19 Vaccines During a Period of XBB.1.5 Dominance in EU/EEA Countries, October to November 2023: A VEBIS-EHR Network Study. Influenza Other Respir Viruses. 2024;18(4):e13292.

11. DeCuir J, Payne AB, Self WH, Rowley EAK, Dascomb K, DeSilva MB, et al.; CDC COVID-19 Vaccine Effectiveness Collaborators. Interim Effectiveness of Updated 2023-2024 (Monovalent XBB.1.5) COVID-19 Vaccines Against COVID-19-Associated Emergency Department and Urgent Care Encounters and Hospitalization Among Immunocompetent Adults Aged 18 Years - VISION and IVY Networks, September 2023-January 2024. MMWR Morb Mortal Wkly Rep. 2024 Feb 29;73(8):180–188.

12. Wells CR, Pandey A, Moghadas SM, Fitzpatrick MC, Singer BH, Galvani AP. Evaluation of Strategies for Transitioning to Annual SARS-CoV-2 Vaccination Campaigns in the United States. Ann Intern Med. 2024;177(5):609–617.

